# Machine Learning to Predict Mortality and Critical Events in COVID-19 Positive New York City Patients

**DOI:** 10.1101/2020.04.26.20073411

**Authors:** Akhil Vaid, Sulaiman Somani, Adam J Russak, Jessica K De Freitas, Fayzan F Chaudhry, Ishan Paranjpe, Kipp W Johnson, Samuel J Lee, Riccardo Miotto, Shan Zhao, Noam D Beckmann, Nidhi Naik, Kodi Arfer, Arash Kia, Prem Timsina, Anuradha Lala, Manish Paranjpe, Patricia Glowe, Eddye Golden, Matteo Danieletto, Manbir Singh, Dara Meyer, Paul F O’Reilly, Laura H Huckins, Patricia Kovatch, Joseph Finkelstein, Robert M Freeman, Edgar Argulian, Andrew Kasarskis, Bethany Percha, Judith A Aberg, Emilia Bagiella, Carol R Horowitz, Barbara Murphy, Eric J Nestler, Eric E Schadt, Judy H Cho, Carlos Cordon-Cardo, Valentin Fuster, Dennis S Charney, David L Reich, Erwin P Bottinger, Matthew A Levin, Jagat Narula, Zahi A Fayad, Allan C Just, Alexander W Charney, Girish N Nadkarni, Benjamin S Glicksberg, on behalf of the Mount Sinai Covid Informatics Center (MSCIC).

**Author notes:** Equal contribution (Sulaiman Somani, Adam J Russak). Correspondence: Allan Just, PhD, Alexander Charney, MD, PhD, Girish Nadkarni, MD, Benjamin Glicksberg, PhD.

## Abstract

Coronavirus 2019 (COVID-19), caused by the SARS-CoV-2 virus, has become the deadliest pandemic in modern history, reaching nearly every country worldwide and overwhelming healthcare institutions. As of April 20, there have been more than 2.4 million confirmed cases with over 160,000 deaths. Extreme case surges coupled with challenges in forecasting the clinical course of affected patients have necessitated thoughtful resource allocation and early identification of high-risk patients. However, effective methods for achieving this are lacking. In this paper, we use electronic health records from over 3,055 New York City confirmed COVID-19 positive patients across five hospitals in the Mount Sinai Health System and present a decision tree-based machine learning model for predicting in-hospital mortality and critical events. This model is first trained on patients from a single hospital and then externally validated on patients from four other hospitals. We achieve strong performance, notably predicting mortality at 1 week with an AUC-ROC of 0.84. Finally, we establish model interpretability by calculating SHAP scores to identify decisive features, including age, inflammatory markers (procalcitonin and LDH), and coagulation parameters (PT, PTT, D-Dimer). To our knowledge, this is one of the first models with external validation to both predict outcomes in COVID-19 patients with strong validation performance and identify key contributors in outcome prediction that may assist clinicians in making effective patient management decisions.

**One-Sentence Summary:** We identify clinical features that robustly predict mortality and critical events in a large cohort of COVID-19 positive patients in New York City.

## Introduction

Despite substantial, organized efforts to prevent disease spread, over 2.4 million people have tested positive for SARS-CoV-2 worldwide, and there have been more than 169,000 deaths to date*(1–3)*. As a result of this pandemic, hospitals are being filled beyond capacity and face extreme challenges with regards to personnel staffing, personal protective equipment availability, and ICU bed allocation. Additionally, patients with COVID-19 demonstrate varying symptomatology, making successful and safe patient triaging difficult. While some infected patients are asymptomatic, others suffer from severe acute respiratory distress syndrome, multiorgan failure, and death*(4)*. Identification of key patient characteristics that govern the course of disease across large patient cohorts is lacking but important, particularly given the potential it has to aid physicians and hospitals in predicting disease trajectory, to allocate essential resources effectively, and to improve patient outcomes. With these needs in mind, we report the development of a decision tree-based machine learning model trained on electronic health records from patients with confirmed COVID-19 status at a single center in the Mount Sinai Health System in New York City to predict critical events and mortality; validate this algorithm at four other hospital centers; and perform a saliency analysis using SHAP (SHapley Additive exPlanation) values to identify the most important features used by this model for outcome prediction.

## Results

### Clinical Data Source and Study Population

We retrieved electronic health records for 3,055 COVID-19-positive inpatient admissions at five hospitals between March 9, 2020 and April 11, 2020 within the Mount Sinai Health System (MSHS). These data included patient demographics, past medical history, and admission vitals and labs (Table 1, Supplementary Table 2). Relevant patient events (intubation, discharge to hospice care, or death) were recorded and subsets were constructed at 3, 5, 7, and 10 day intervals after admission (Figure 1). Of these patients, 17.0% to 31.6% had a critical event (intubation, discharge to hospice care, or death) and 6.0% to 21.5% died over the observed time frames (Supplementary Table 1). In contrast, the control group consisted of patients with all other discharge dispositions and those that were still hospitalized.

**Table 1.** Characteristics of Hospitalized Covid-19 Patients at Baseline (n= 3055)

| Characteristics of Admission | MSH | Other locations |
| --- | --- | --- |
| **Demographics** |  |  |
| **Sex, n (%)** |  |  |
| Male | 704 (57.5) | 1070 (58.5) |
| Female | 521 (42.5) | 760 (41.5) |
| **Race, n (%)** |  |  |
| Other | 537 (43.8) | 749 (40.9) |
| Caucasian | 293 (23.9) | 435 (23.8) |
| African American | 273 (22.3) | 503 (27.5) |
| Unknown | 64 (5.2) | 60 (3.3) |
| Asian | 44 (3.6) | 65 (3.5) |
| Pacific Islander | 14 (1.1) | 18 (0.9) |
| **Ethnicity, n (%)** |  |  |
| Non-Hispanic Latino | 676 (55.2) | 1133 (61.9) |
| Hispanic/Latino | 333 (27.2) | 503 (27.5) |
| Unknown | 216 (17.6) | 194 (10.6) |
| **Age, Median (IQR)** | **62.1 (49.4-71.8)** | **68.33 (56.9-78.8)** |
| **Age group, n (%)** |  |  |
| 18-30 | 56 (4.6) | 37 (2.0) |
| 31-40 | 139 (11.6) | 93 (5.1) |
| 41-50 | 133 (10.9) | 158 (8.6) |
| 51-60 | 242 (19.8) | 309 (16.9) |
| 61-70 | 320 (26.1) | 431 (23.5) |
| 71-80 | 202 (16.5) | 422 (23.1) |
| 81-90 | 102 (8.3) | 308 (16.8) |
| At least 90 | 31 (2.5) | 72 (3.9) |
| **Previous Medical History, n (%)** |  |  |
| Hypertension (%) | 462 (37.7) | 686 (37.5) |
| Atrial Fibrillation (%) | 86 (7.0) | 130 (7.1) |
| Coronary Artery Disease (%) | 171 (13.9) | 307 (16.8) |
| Heart failure (%) | 110 (8.9) | 181 (9.9) |
| Stroke (%) | 98 (8) | 118 (6.5) |
| Chronic Kidney Disease (%) | 136 (11.1) | 158 (8.6) |
| Diabetes (%) | 313 (25.5) | 466 (25.5) |
| Asthma (%) | 115 (9.4) | 132 (7.2) |
| Chronic Obstructive Pulmonary Disease (%) | 65 (5.3) | 103 (5.6) |
| Cancer (%) | 112 (9.1) | 94 (5.1) |
| **Vital Signs at Hospital Admission, Median (IQR)** |  |  |
| Heart Rate (bpm) | 89 (78 -100) | 89 (78 - 100) |
| Pulse Oximetry (%) | 96 (94 - 98) | 96 (94 - 98) |
| Respiration Rate (breaths / minute) | 20 (18 - 20) | 18 (18 - 20) |
| Temperature (F) | 98.7 (98.1 - 99.9) | 97.9 (98.6 - 99.5) |
| Systolic Blood Pressure (mmHg) | 124 (112 - 138) | 127 (112 - 142) |
| Diastolic Blood Pressure (mmHg) | 69 (61 - 78) | 72 (65 - 81) |
| Weight (kg) | 80.9 (68.9 - 95.3) | 78.9 (68.04 - 91.7) |
| **Admission Laboratory Parameters, Median (IQR)** |  |  |
| **Metabolic markers** |  |  |
| Sodium (mEq/L) | 137 (135 - 140) | 138 (135 -141) |
| Potassium (mEq/L) | 4 (3.6 - 4.5) | 4.2 (3.9 - 4.7) |
| Creatinine (mg/dL) | 0.9 (0.7 - 1.4) | 1.0 (0.8 -1.6) |
| Lactate (mg/dL) | 1.7 (1.3 - 2.2) | 1.4 (1.1 - 2.0) |
| **Hematological markers** |  |  |
| White Blood Cells ($10^3/\mu\text{L}$) | 6.8 (5.2 - 9.8) | 7.7 (5.7 - 10.5) |
| Lymphocyte Percentage | 13.1 (7.8 - 20.7) | 14.1 (8.9 - 21.4) |
| Hemoglobin (mEq/L) | 12.5 (11.1 - 13.7) | 12.8 (11.4 - 14) |
| Red Blood Cell Distribution Width (%) | 14.4 (13.6 -15.6) | 12.7 (12.1 - 13.9) |
| Platelets (#) | 212 (162 - 279.3) | 194 (148 - 260) |
| **Liver Function** |  |  |
| Alanine Aminotransferase (units/L) | 31 (19 - 54) | 31 (20 - 53) |
| Aspartate Aminotransferase (units/L) | 41.5 (29 - 67) | 47 (31 - 74) |
| Albumin (g/dL) | 3 (2.6 - 3.3) | 3 (2.6 - 3.3) |
| Total Bilirubin (mg/dL) | 0.6 (0.4 - 0.8) | 0.6 (0.4 - 0.8) |
| **Coagulation markers** |  |  |
| Prothrombin Time (s) | 14.3 (13.5 - 15.7) | 14.2 (13.5 - 15.5) |
| Partial Thromboplastin Time (s) | 32.5 (29.2 - 37.1) | 33.3 (30 - 38.8) |

| **Gases** |  |  |
| --- | --- | --- |
| PCO<sub>2</sub> (mmHg) | 37 (41 - 47) | 41 (35 - 47) |
| pH | 7.4 (7.4 - 7.4) | 7.4 (7.3 - 7.4) |
| **Inflammatory markers** |  |  |
| C Reactive Protein (mg/L) | 152.3 (83.1 - 205.6) | 124.6 (62.3 - 214.5) |
| Ferritin (ng/mL) | 720 (343.5 - 1813.8) | 818 (388 - 1966) |
| D-Dimer (ng/mL) | 1.3 (0.7 - 2.5) | 1.5 (0.9 - 3.0) |
| Creatinine Phosphokinase (units/L) | 156 (80 - 505) | 237.5 (187.8 - 440.5) |
| Lactate Dehydrogenase (units/L) | 417 (315 - 559) | 450 (346.3 - 610.8) |
| Procalcitonin (ng/mL) | 0.2 (0.1 - 0.6) | 0.2(0.1 - 0.7) |
| **Cardiac markers** |  |  |
| Troponin I (ng/mL) | 0.1 (0.02 - 0.2) | 0.2 (0.1- 0.9) |

**Fig 1.**
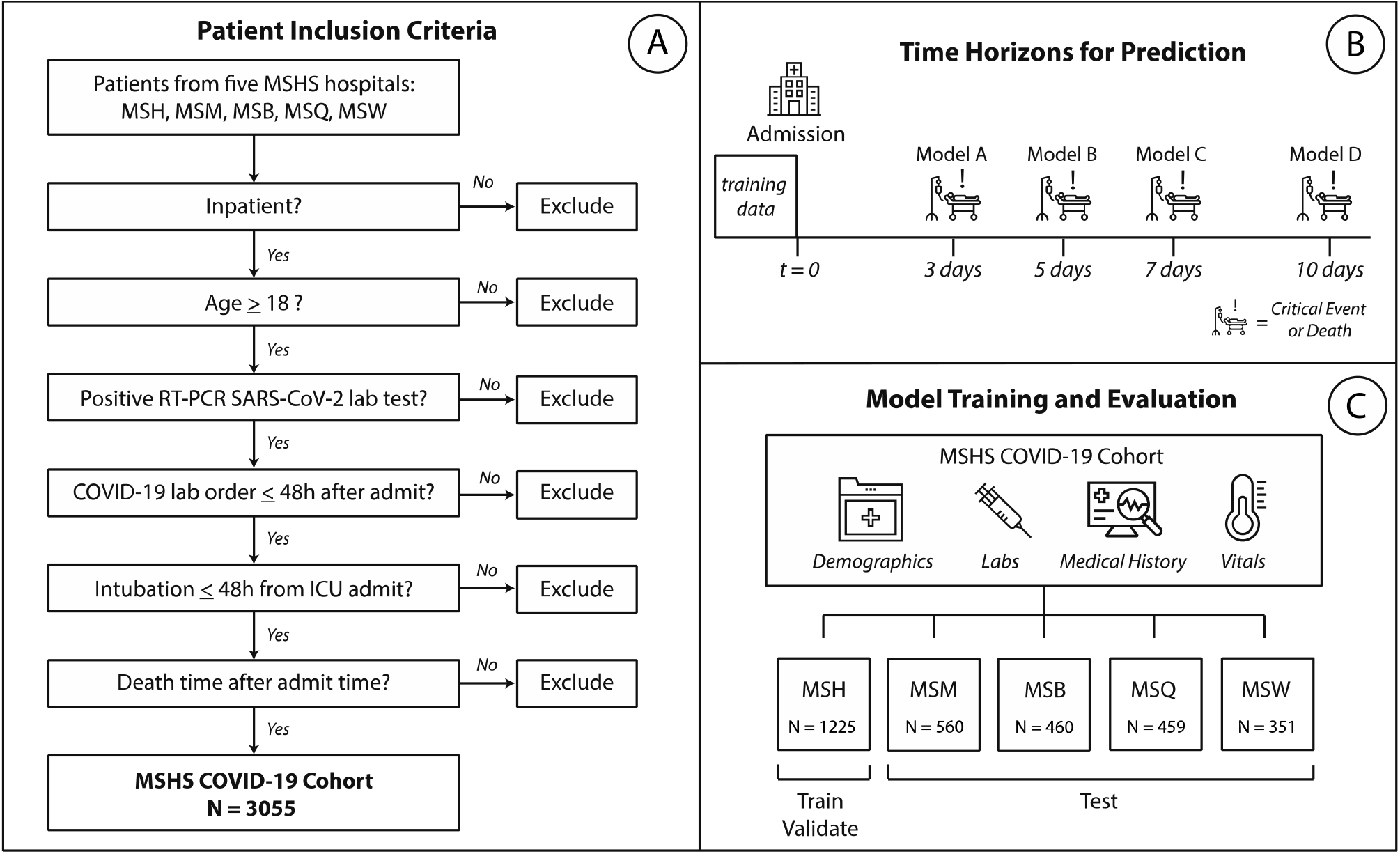
Study Design and Workflow. **A)** Procedure for patient inclusion in our study. **B)** Strategy and design for experiments. Patient clinical data from Mount Sinai Hospital (MSH) was used to train and validate our machine learning model. We then test these models on patients from four other external hospitals within the Mount Sinai Health System: Mount Sinai Brooklyn (MSB), Mount Sinai Morningside (MSM), Mount Sinai Queens (MSQ), and Mount Sinai West (MSW). **C)** Machine learning experimental design. We train on data from admission to predict mortality and critical illness outcomes.

### Classifier training and performance

Given the large number of patients in the analysis and presence of missing variables in the data, we used XGBoost*(5)*, a boosted decision-tree based machine learning (ML) model, to predict either a critical event or death of a patient within the aforementioned time frames. Patients from the Mount Sinai Hospital (MSH) were split into a training and validation set for the model. To increase model generalizability and help minimize bias, the model’s performance was assessed on a test set composed entirely of patients from the other hospitals (OH) in the MSHS. While multiple time limits for event occurrence were assessed, the results and discussion in this letter focus predominantly one week after admission. As a control, both simple and generalized additive logistic regression models were trained to assess performance, given their ubiquity as the preferred model in current COVID research pieces.

After training, the classifier robustly predicted the presence of a critical event at three, five, seven, and 10 days (Figure 2 and Supplementary Table 4) as measured by area under the receiver operating characteristic curve (AUC-ROC = 0.74 for OH, 0.83 for MSH at 1 week) and area under the precision-recall curve (AUPRC = 0.58 for OH, 0.49 for MSH at 1 week). We were able to achieve similar performance at predicting critical events at longer intervals, namely 15 and 20 days (Supplementary Table 5). As a baseline comparison, the logistic regression models (Supplementary Table 6) performed more poorly in prediction by AUC-ROC (0.52 - 0.74) and AUC-PRC (0.19 - 0.37) for critical events at three days relative to the XGBoost model (AUC-ROC = 0.77, AUC-PRC = 0.43). With respect to mortality, the model achieved high specificity (0.79 - 0.92 for OH) and AUC-ROC (0.79 - 0.92 for OH) with similar AUPRC (0.38 - 0.65 OH) as with critical events. As the event time window increased, the performance of the classifier by the AUPRC value improved, which likely stems from the infrequency of deaths at earlier time points that created a class imbalance for mortality. Comparatively, all logistic regression models underperformed significantly in prediction by AUC-ROC (0.61-0.70) and AUC-PRC (0.06 - 0.18) for mortality.

**Fig 2.**
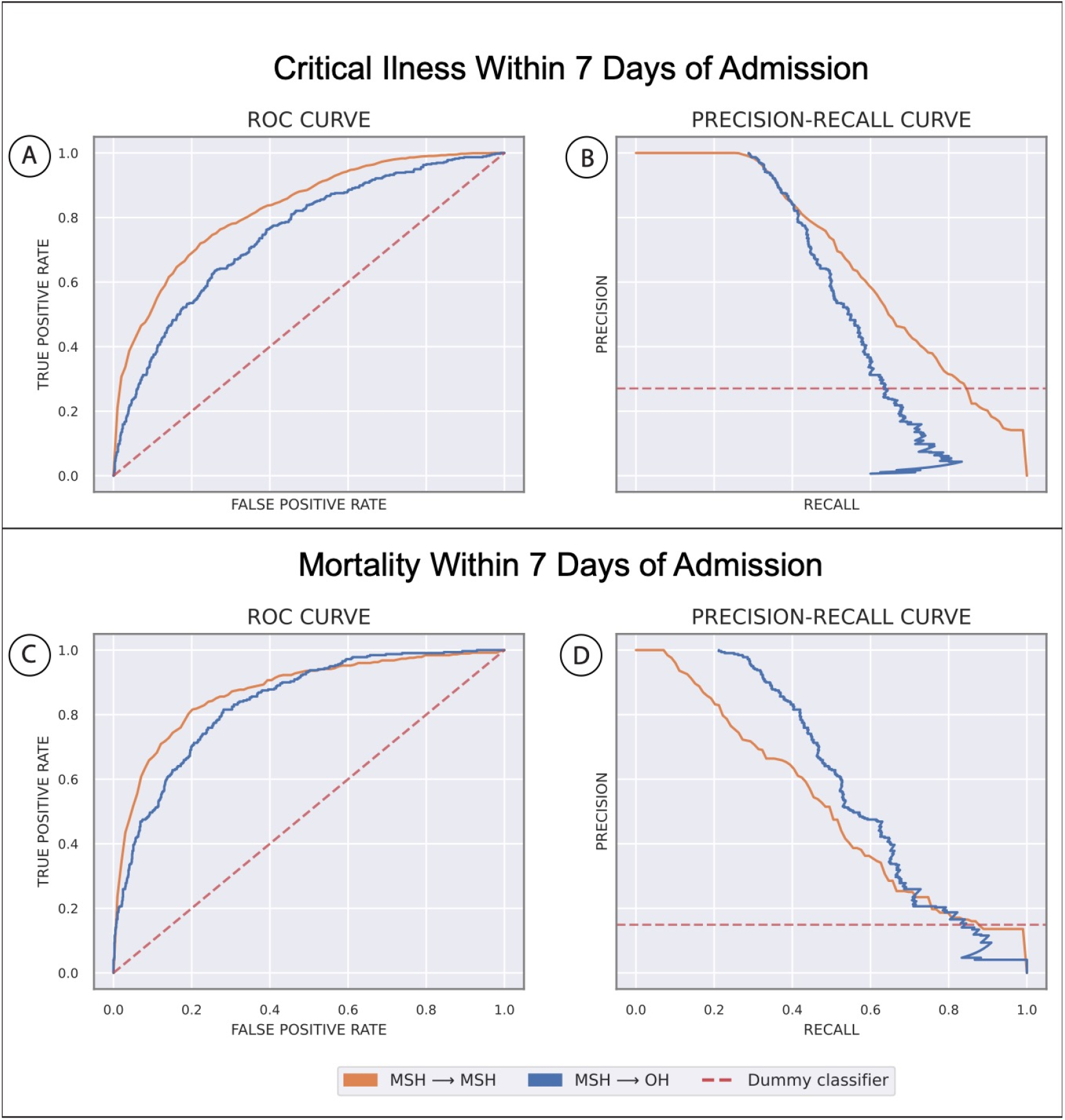
Machine learning experimental results. In all plots, the orange line reflects training at Mount Sinai Hospital (MSH) and testing via cross-validation. The blue line reflects testing the model built on patient data from MSH on external patients from all other hospitals (OH) **A)** Area under the receiver operator characteristic curve (AUC-ROC) for predicting critical illness at seven days since admission. **B)** Area under the precision-recall curves (AUPRC) for predicting critical illness at seven days since admission. **C)** AUC for predicting mortality at seven days since admission. **D)** AUPRC for predicting critical illness at seven days since admission.

### Identifying important features in the model

To identify the most salient features driving model prediction, SHAP (SHapley Additive exPlanations) values*(6)* were calculated for the highest-performing model in the cross-validation set during hyperparameter tuning (Figure 3). Among the top features, both high and low levels of lactate dehydrogenase (LDH), procalcitonin, and D-dimer were strong drivers for predicting a critical event at 1 week, while elevated prothrombin time (PT) and partial thromboplastin time (PTT) favored the classifier to predict a critical event. For mortality, both high and low values for age, procalcitonin, and red blood cell distribution width (RDW) were the strongest effectors in guiding mortality prediction by the model within 1 week of admission. Other important variables for increasing the prediction for death included an elevated troponin, LDH, lymphopenia (i.e. low lymphocyte percentage), white blood cell count (WBC), aspartate aminotransferase (AST), and D-dimer. Finally, using SHAP interaction scores, we discovered that covariate interactions between features, relative to each feature’s independent importance, contributed less to the model’s prediction (Supplementary Figures 1-4).

**Fig 3.**
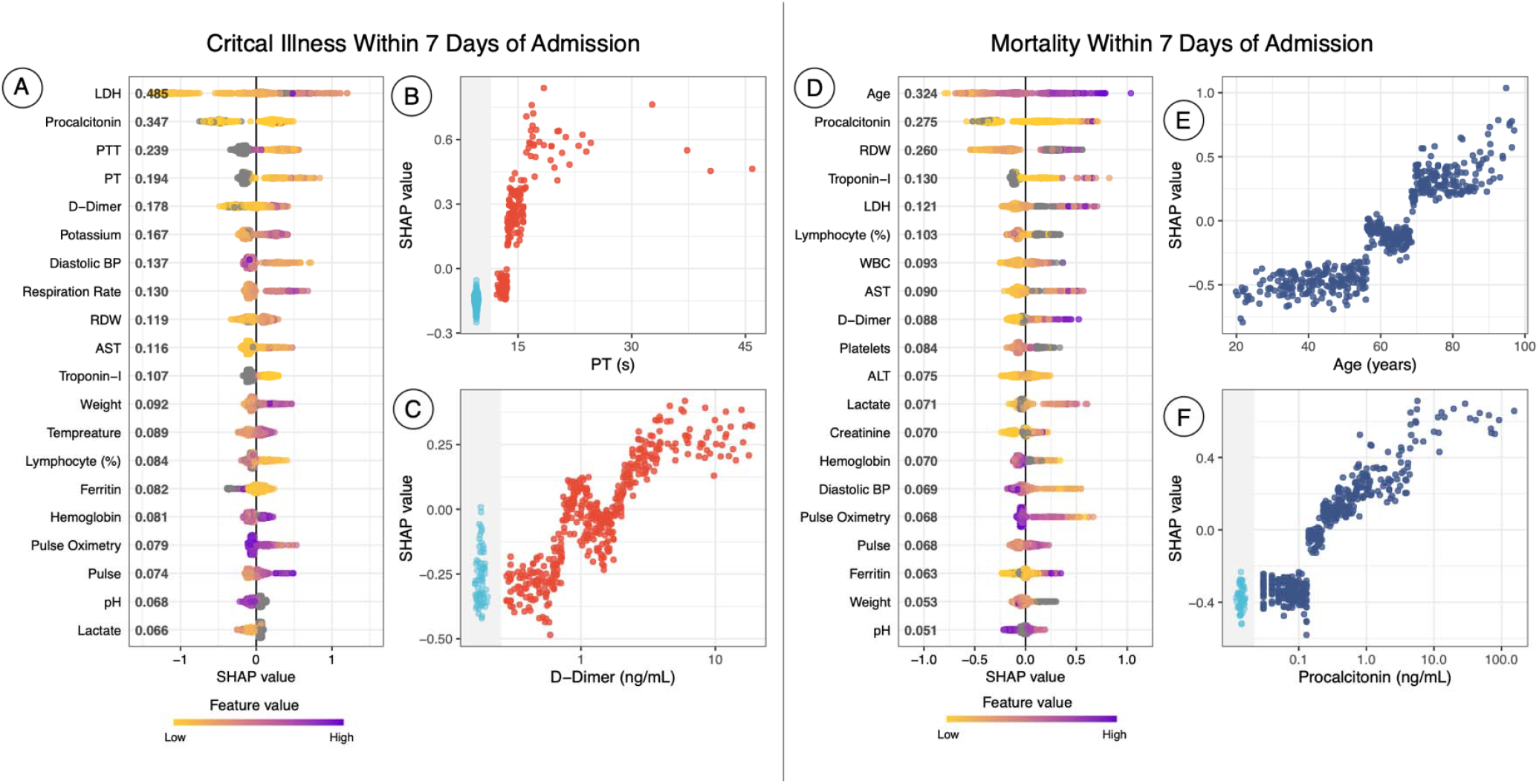
SHAP Summary and Dependency Plots. SHAP summary plots for critical event **(A)** and mortality **(D)** at 7 days show the SHAP values for the most important features for the respective XGBoost model. Features in the summary plots (y-axis) are organized by their mean absolute SHAP values (x-axis), which represents the importance of that feature in driving the classifier’s prediction, for patients. Values of those features for each patient (i.e. a particular LDH value) are colored by their relative value. **(B)** and **(C)** represent dependency plots, which similarly demonstrate how different values of those features can affect the SHAP score and ultimately impact classifier decisions, for prothrombin time (PT) and D-Dimer, respectively, for critical event prediction. **(E)** and **(F)** represent dependency plots for age and procalcitonin levels. Patients with missing values for a feature in the dependency plot are clustered in the shaded area to the left.

## Discussion

We highlight several important findings that have implications in clinical medicine. First, we offer robust prediction algorithms pertaining to the most clinically severe outcomes based solely on admission metrics. This insight provides the likely hospital course for patients up to 10 days into the future. High sensitivity in predicting mortality within three, five, and seven days of admission (0.86 - 1.0) suggests that this model can potentially be used by clinicians in gauging the acute clinical course of an admitted patient. The model’s high specificity, particularly for mortality at days three (specificity = 0.92) and five (specificity = 0.86), suggest its role for augmenting clinicians’ decision-making when identifying patients at immediate risk of impending clinical decompensation and potentially guide allocation of more intensive care upon admission.

Additionally, our framework permits a clinically relevant understanding of the model’s most salient features defining its decision boundaries. Age was the most important feature for mortality prediction in COVID-19+ patients, with a notable exponential rise of feature contribution as age increased (Figure 3)*(7, 8)*. Elevations in serum LDH, although nonspecific markers of inflammation, are implicated in pulmonary endothelial cell injury and in COVID-19+ patients*(9–11)*. Equivalently, procalcitonin has been implicated as a biomarker of underlying infection and sepsis risk*(12, 13)*. Elevated RDW, which may be an index for enhanced patient frailty and risk of adverse outcomes*(14)*, was also a strong driver of mortality. Other salient features like leukocytosis*(15)*, a natural response to inflammation, in combination with virally-driven lymphopenia*(16, 17)*, have also been associated with COVID-19 burden. Additionally, elevated troponins*(18, 19)*, renal dysfunction from elevated creatinine*(20)*, anemia, vital instability (low oxygen saturation, tachycardia, hypotension), elevated ferritin*(10, 19)*, high lactate, and acidosis were also contributors to driving model prediction towards mortality. With growing evidence of COVID-19-induced hypercoagulable states in these patients*(10, 21, 22)*, it is promising that our model recognized the feature importance of coagulability markers, such as PT, PTT, and D-dimer (Figure 3). Thus, this corroboration of the features learned by XGBoost and highlighted by the SHAP analysis with those findings from pathophysiological principles and more recent correlative studies exploring COVID-19 patients*(2, 3, 18, 23–25)* gives additional credibility to these findings.

Just as interesting as the features present in the SHAP value analysis were those that were not. For example, because race is both poorly represented (“Unknown”) and categorized inadequately in electronic health records, the model did not find race to be important for outcome prediction and instead opted to favor more objective data (vitals, labs). Contrary to our expectation, age was not identified as a significant feature for critical event prediction in these primary analyses. However, SHAP value analyses for critical event prediction at longer time frames (10, 15, and 20 days) revealed an increasing importance on age for outcome prediction (Supplementary Figure 5). This trend suggests the model’s decision to capture acute events by relying on more objective measures not confounded by other factors that are cached into age, which may better represent illness severity. However, over time, age may become a better marker for critical event prediction, by offering a more stable container of clinical information, given its invariance to change relative to other features. As such, the classifier becomes optimized for predicting severe events earlier in the course of illness.

The results of our models should be considered in light of several limitations. First, we base our predictions solely on a patient’s admission labs (i.e. within 36 hours); while this restriction encourages the use of this model in patient triage, events during a patient’s hospital stay after admission may drive their clinical course away from the prior probability. Furthermore, not all patient labs are drawn at admission, which introduces an element of missingness in our dataset. For example, unlike the general patient population, patients on anticoagulation therapy, who likely have comorbidities increasing their baseline risk, will have coagulation labs (PT, PTT) taken on admission. However, the shift away from predicting death by the model in the absence of PT/PTT (Figure 3) suggests that missingness in coagulation labs is a proxy for this lower baseline risk secondary to not having comorbid conditions that require anticoagulation therapy. Additionally, patients admitted to the hospital later in the crisis were both beneficiaries of improved patient care protocols from experiential learning, but also victims of resource constraints from overburdened hospitals. These effects, while possibly negated by our large sample size, may also induce temporal variation between patient outcomes. Furthermore, inherent limitations exist when using EHRs, especially those integrated from multiple hospitals. In order to facilitate timely dissemination of our results, we chose not to manually chart review patient notes that may have otherwise provided additional potential features, such as symptoms and clinical course, to incorporate in our model. Because all five hospitals operate in a single health system, system-wide protocols in lab order sets and management protocols were an additional source of bias that may lower external validity. Other interhospital effects such as shuttling COVID-19 cases to certain hospitals for balancing systemic patient burden may also imbalance case severity across hospitals and care management between hospitals; certainly, this was the case for MSW, where mortality at 3-days was far lower (1.7%) than other hospital sites. This was ultimately a major reason to restrict model training to a single center and perform testing out of sample in another hospital center. Finally, though XGBoost is superior to other models at handling missing data, a notable drawback is its bias towards continuous features instead of categorical ones, given increased information represented in the form*(28)*. However, collinearities between some categorical features in this dataset may be present with other continuous features, as exhibited by covariance strength between hypertension and systolic BP and creatinine in Supplemental Figure 4, which can then serve as vehicles for capturing these categorical pieces of information.

In conclusion, the COVID-19 pandemic unequivocally represents an unprecedented public health crisis. Healthcare institutions are facing extreme difficulties in managing resources and personnel. Physicians are treating record numbers of patients and continuously expose themselves to a highly contagious and virulent disease with varying symptomatology. Only few therapeutic options have demonstrated improvement to patient outcomes. As COVID-19 moves outside of the current epicenter in New York City, healthcare institutions will see a larger influx of affected patients and can benefit from immediate insights regarding assessment of disease severity*(29, 30)*. These models successfully predict critical illness and mortality up to 10 days in advance in a diverse patient population from admission information alone and provide important markers for acute care prognosis that can be used by healthcare institutions to improve care decisions at both the physician and hospital level for management of COVID-19 positive patients.

## Materials and Methods

This study has been approved by the Institutional Review Board at the Icahn School of Medicine at Mount Sinai (IRB-20-03271).

### Clinical Data Source and Study Population

In this study, patient data came from five hospitals within the Mount Sinai Hospital System (MSHS): the Mount Sinai Hospital (MSH) located in East Harlem, Manhattan; Mount Sinai Morningside (MSM) located in Morningside Heights, Manhattan; Mount Sinai West (MSW) located in Midtown and the West Side, Manhattan; Mount Sinai Brooklyn (MSB) located in Midwood, Brooklyn; and Mount Sinai Queens (MSQ) located in Astoria, Queens. The dataset was obtained from different sources and aggregated by the Mount Sinai COVID Informatics Center (MSCIC).

We included patients who were over 18 years old that had a laboratory-confirmed COVID-19 infection, and were admitted between March 9 and April 11, 2020 to any of the hospitals previously mentioned. A confirmed case of COVID-19 was defined by a positive reverse transcriptase polymerase chain reaction (RT-PCR) assay of a nasopharyngeal swab. We excluded patients who had a positive COVID-19 RT-PCR result more than two days after admission. Additional exclusion criteria are presented in Figure 1. Full patient characteristics by site are provided in Supplementary Table 1.

### Study Data

Demographics included age, sex, as well as reported race, and ethnicity. Race was collapsed into seven categories based off of the most recent US census race categories: American Indian or Alaskan Native, Asian, Black or African-American, Other, Native Hawaiian or Other Pacific Islander, Unknown, and White. Ethnicity was collapsed into three categories: Hispanic/Latino, Non-Hispanic/Latino, and Unknown. We obtained demographics, diagnosis codes (International Classification of Diseases-9/10-Clinical Modification (ICD-9/10-CM) codes and procedures), as well as vital signs and laboratory measurements during hospitalization. A pre-existing condition was defined as the presence of ICD-9/10-CM codes associated with specific diseases. We chose to include as covariates conditions that have been previously reported to have increased incidence in hospitalized COVID-19 patients, specifically: atrial fibrillation, asthma, cancer, coronary artery disease, chronic kidney disease, chronic obstructive pulmonary disease, diabetes mellitus, heart failure, hypertension, and stroke*(15, 23–25, 31)*.

We included laboratory measurements and vital signs near the time of admission for prediction. Specifically, because records for laboratory values and vitals may appear with some lag, only the first available value within 36 hours of admission was included, otherwise the value was assigned as missing. Height was absent in a large percentage of the patients (18.2%). Because height is generally invariant in the adult population, and given the resource constraint of the pandemic, it was common for triage nurses to use the height from a previous and recent admission. In an effort to be as cohesive in our data gathering methods as possible, these earlier records were not retrieved for this dataset. However, weight was used as the next approximate proxy for body habitus, with additional information being presented through sex and age for body habitus as well.

All lab orders from the five hospitals were queried for patients included in this study within the timeframe of interest. Due to discrepancies in how labs were named in different hospitals, a comprehensive review of all lab field names was conducted by a multidisciplinary team of clinical and statistical experts to ensure that there was a direct mapping between all sites. Additionally, many labs represented a single component (e.g. sodium), but were acquired from either an arterial blood gas (ABG), venous blood gas (VBG), and basic metabolic panel (BMP). Based on the utility of these lab values in clinical practice and the similarity between their statistical distributions, labs derived from a VBG or BMP were collapsed into a single category (i.e. ‘SODIUM’) and those derived from an ABG were moved to a separate category (i.e. ‘SODIUM_A’). Finally, the earliest lab, by time of result, in the set of all lab order names that were combined into a single lab category was chosen as the representative lab value for that category. Finally, lab data below the 0.5th and above the 99.5th percentiles were removed to avoid inclusion of any obvious outliers that could represent incorrect documentation and measurement error.

### Definition of Outcomes

The two primary outcomes were 1) death versus survival or discharge, and 2) critical illness versus survival or discharge, through time horizons of 3, 5, 7, and 10 days. Critical illness is defined as discharge to hospice, intubation ≤ 48 hours prior to ICU admission, or death. To address potential concerns of censoring by limiting exploration of only these time frames, particularly in the case of model enrichment for acute critical events, we also predicted critical events at days 15 and 20.

### Statistical Analysis

Our primary model was fit with the Extreme Gradient Boosting (XGBoost) implementation of boosted decision trees on continuous and one-hot encoded categorical features. The XGBoost algorithm provides state-of-the-art prediction results through an iterative process of averaging in decision trees (we used 100) fit to the residual error of the prior ensemble. While each tree is too simple to accurately capture complex phenomena, the combination of many trees in the XGBoost model accommodates non-linearity and interactions between predictors. Missing data values are routed through split points based on the direction to minimize loss. XGBoost models were trained and evaluated using 10-fold stratified cross validation. For each fold, hyperparameter tuning was performed by randomized grid searching directed towards maximizing the sensitivity metric over 2,000 discrete grid options. Cross-validation was performed inside each grid option. We present the model hyperparameters for all experiments in Supplementary Table 3. The performance of the models were measured using the area under the receiver operator characteristic curve (AUC-ROC), area under the precision-recall curve (AUC-PRC), F1-score, sensitivity, and specificity. To interpret the significance of input features on the model’s prediction, SHAP values across all features on the best-performing model, by AUC-ROC, in the cross-validation set were calculated. Finally, we tested these models built on patient data from MSH on patients from four other hospitals (Figure 1).

As a baseline, we also fit a logistic regression model and a generalized additive logistic model to compare our XGBoost model. We decided to use a generalized additive model because of its ability to extend generalized linear models by allowing for non-linear functions of features. Four main models were generated: 1) Logistic regression using only Age 2) Generalized Additive Model using only Age 3) Logistic Regression with all available features 4) Generalized Additive Model using all available features. Since these models do not have a built in method of dealing with missing data, we dropped features with over 70% missingness and samples that lacked values for the remaining feature space. The models were trained and evaluated using 10-fold stratified cross validation on patient data from MSH and subsequently evaluated on patient data from the other hospitals. The same metrics were recorded for these models; however, this model was only trained at outcome prediction on Day 3, which was the time frame at which model AUC-ROC was highest for the XGBoost classifier. Performance results for this classifier are represented in Supplementary Table 5.

## Data Availability

n/a

---

**Supplementary Fig 1.**
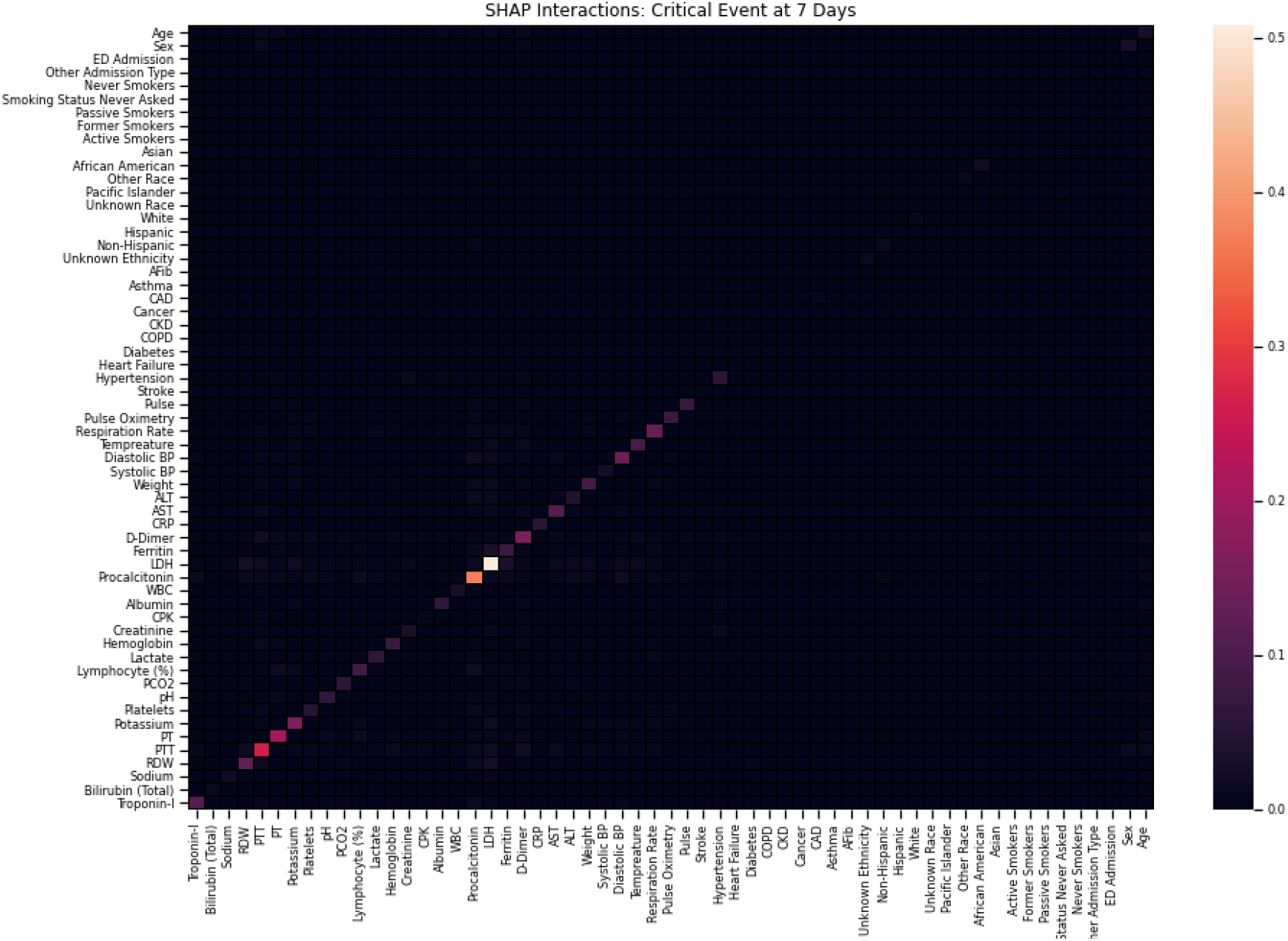
SHAP Interactions: Critical Event at 7 Days. Heatmap demonstrating the composite SHAP interaction scores between features for patients from the best performing k th validation fold for critical event prediction at 7 days. The intensity along the diagonal represents independent contributions of a feature towards model prediction (i.e. mean absolute SHAP values in Figure 3). Covariant interactions between features is significantly less relative to the intensity of the independent contributions of each feature towards model prediction.

**Supplementary Fig 2.**
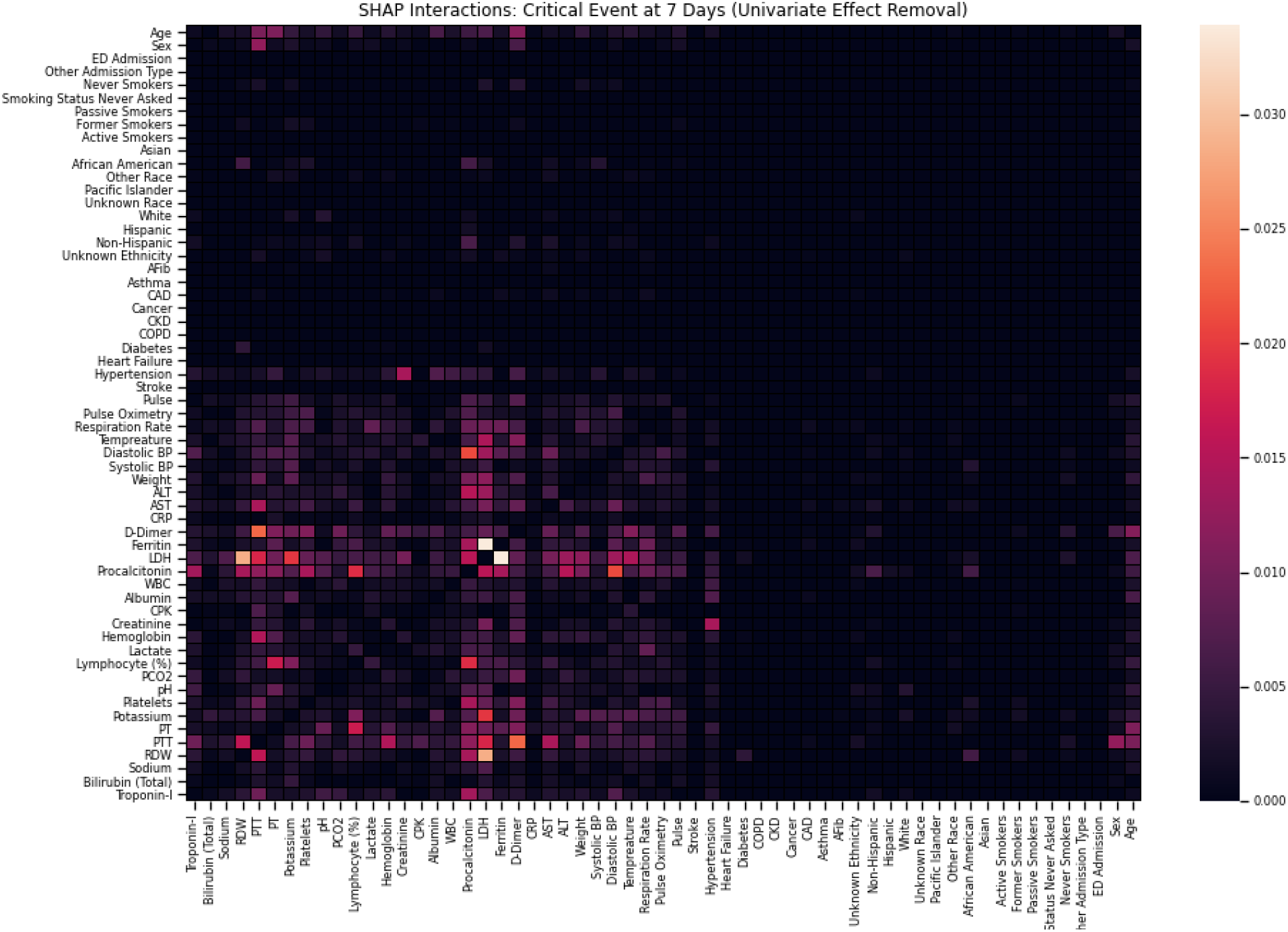
SHAP Interactions: Critical Event at 7 Days (Univariate Effect Removal). Heatmap demonstrating the composite SHAP interaction scores between features for patients from the best performing k th validation fold for critical event prediction at 7 days. Univariate feature contributions along the diagonal have been set to 0 to better examine the relative strength of covariance between features.

**Supplementary Fig 3.**
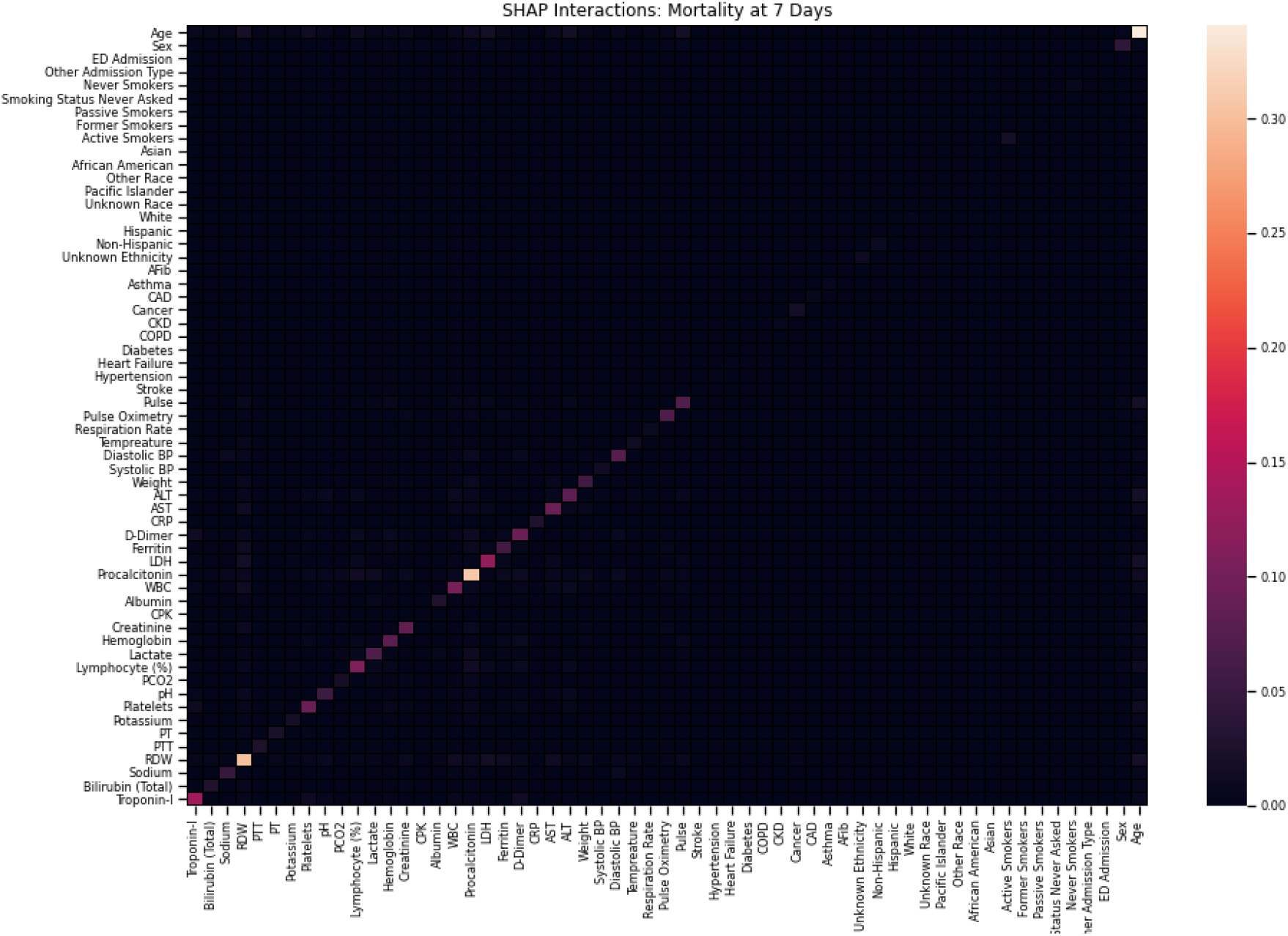
SHAP Interactions: Mortality at 7 Days. Heatmap demonstrating the composite SHAP interaction scores between features for patients from the best performing k th validation fold for mortality prediction at 7 days. The intensity along the diagonal represents independent contributions of a feature towards model prediction (i.e. mean absolute SHAP values in Figure 3). Covariant interactions between features is significantly less relative to the intensity of the independent contributions of each feature towards model prediction.

**Supplementary Fig 4.**
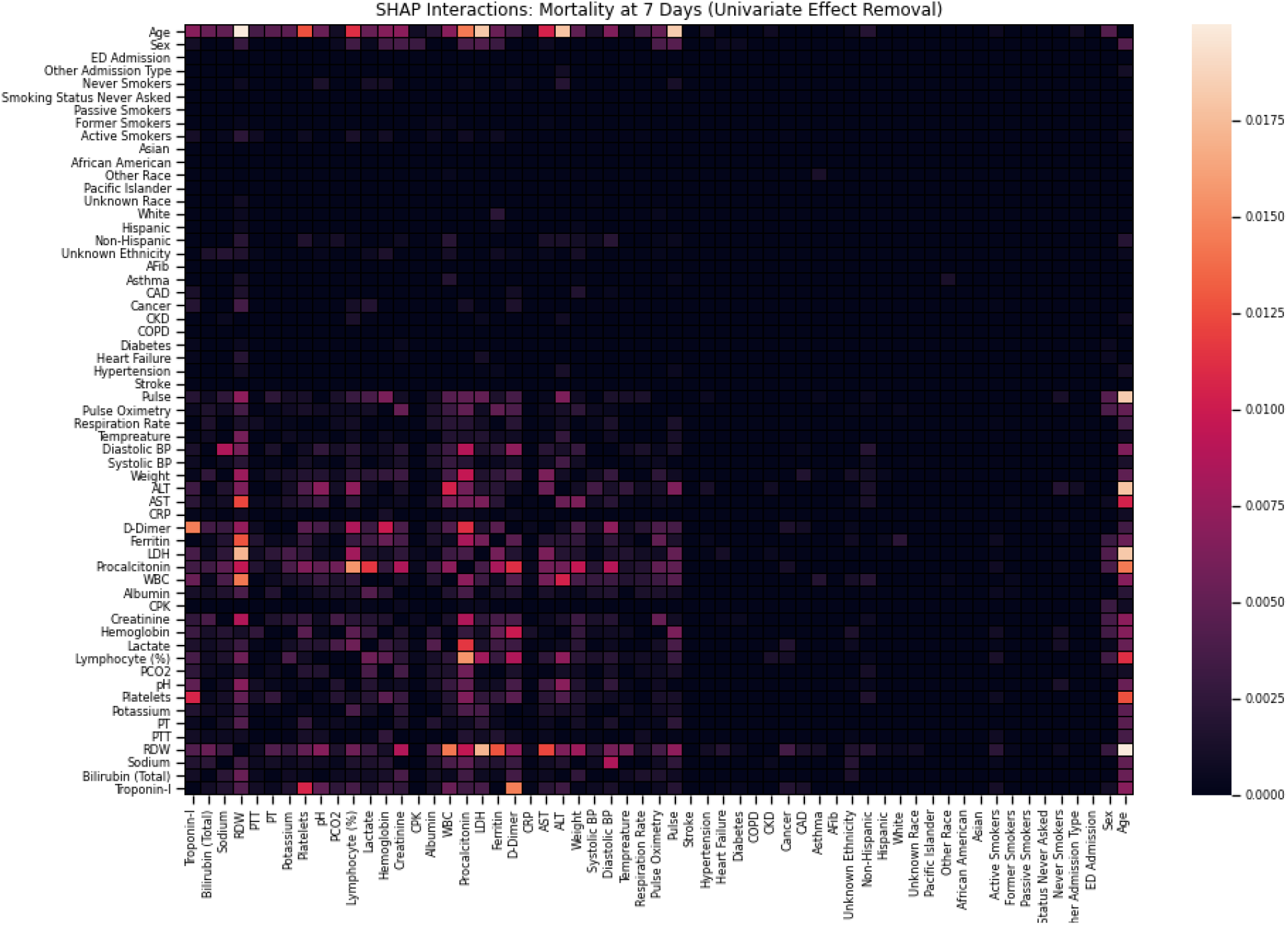
SHAP Interactions: Mortality at 7 Days (Univariate Effect Removal). Description: Heatmap demonstrating the composite SHAP interaction scores between features for patients from the best performing k-th validation fold for mortality prediction at 7 days. Univariate feature contributions along the diagonal have been set to 0 to better examine the relative strength of covariance between features.

**Supplementary Fig 5.**
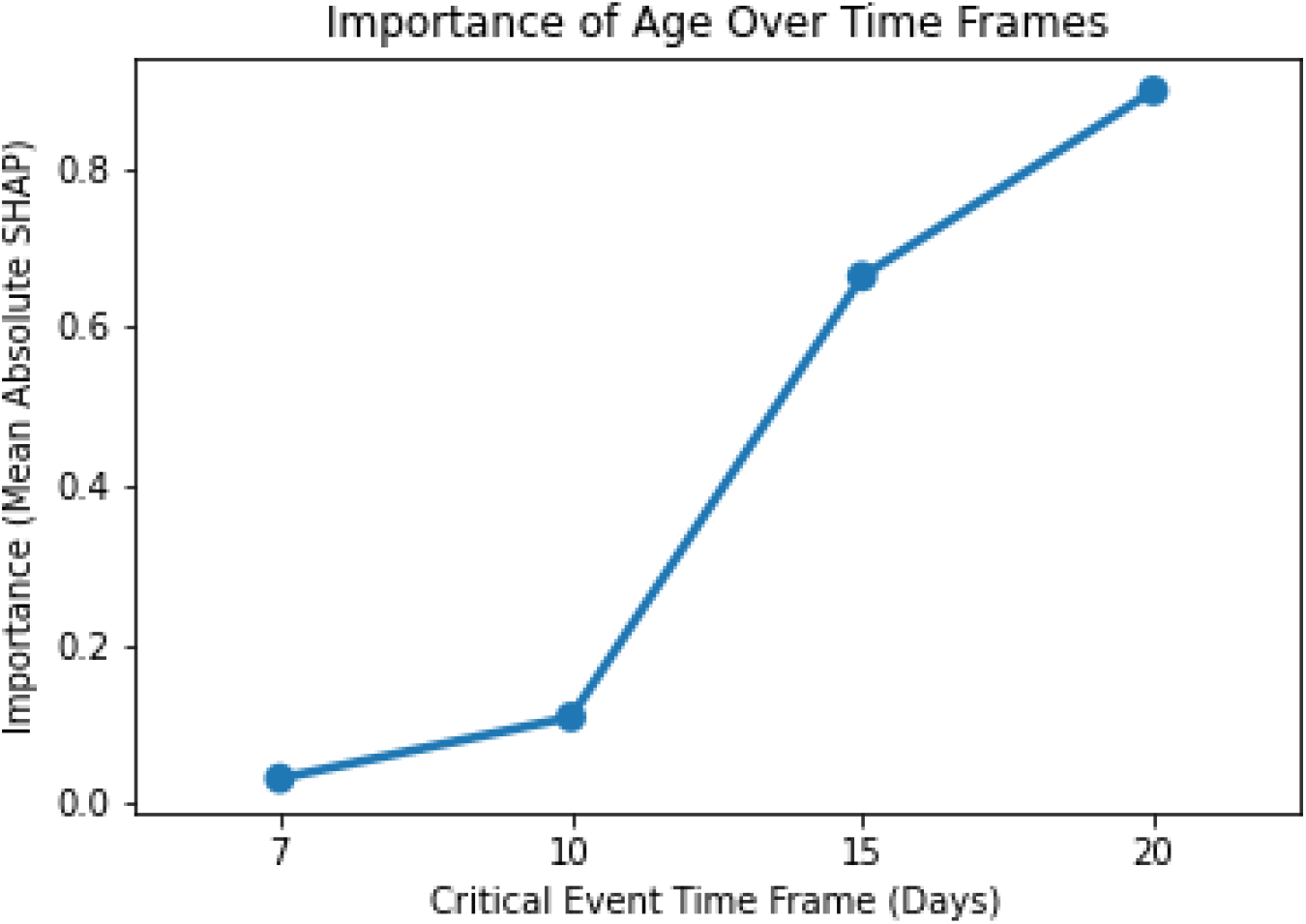
Importance of Age Over Time Frames. Mean absolute SHAP value for age (blue) to represent importance in prediction of critical event occurrence over different time frames (7, 10, 15, and 20 days).

**Supplementary Table 1.** Baseline Patient Characteristics by Hospital. Characteristics, such demographics, clinical history, vital signs, and laboratory tests, for all patients included in the study and delineated by the hospital site at which the patient was admitted.

| Characteristics or Admission | MSQ | MSB | MSH | MSW | MSM |
| --- | --- | --- | --- | --- | --- |
| **Demographics** |  |  |  |  |  |
| **Sex, n (%)** |  |  |  |  |  |
| Male | 291 (63.4) | 207 (58.9) | 317 (56.6) | 255 (55.4) | 704 (57.47) |
| Female | 168 (36.6) | 144 (41.0) | 243 (43.4) | 205 (44.6) | 521 (42.53) |
| **Race, n (%)** |  |  |  |  |  |
| Other | 33 (7.2) | 9 (1.9) | 44 (3.6) | 16 (4.6) | 7 (1.3) |
| African American | 50 (10.9) | 191 (41.5) | 273 (22.3) | 68 (19.4) | 194 (34.6) |
| Caucasian | 264 (57.5) | 91 (19.8) | 537 (43.8) | 131 (37.3) | 263 (46.9) |
| Asian | 9 (1.9) | 0 (0) | 14 (1.1) | 4 (1.1) | 5 (0.9) |
| Pacific Islander | 16 (3.5) | 19 (4.1) | 64 (5.2) | 5 (1.4) | 20 (3.6) |
| Unknown | 87 (18.9) | 150 (32.6) | 293 (23.9) | 127 (36.2) | 71 (12.7) |
| **Ethnicity, n (%)** |  |  |  |  |  |
| Hispanic/Latino | 230 (50.1) | 322 (70) | 676 (55.2) | 251 (71.5) | 330 (58.9) |
| Non-Hispanic Latino | 188 (40.9) | 17 (3.7) | 333 (27.2) | 92 (26.2) | 206 (36.8) |
| Unknown | 41 (8.9) | 121 (26.3) | 216 (17.6) | 8 (2.3) | 24 (4.3) |
| **Age - Median(IQR)** | 66 (56-77) | 65.3 (51.7-76.4) | 68.8 (56.0-80.1) | 72 (63-82) | 62.1 (49.4-71.8) |
| **Age group, n (%)** |  |  |  |  |  |
| 18-30 | 10 (2.2) | 16 (4.6) | 7 (1.3) | 4 (0.9) | 56 (4.6) |
| 31-40 | 16 (3.5) | 38 (10.8) | 32 (5.7) | 7 (1.5) | 139 (11.4) |
| 41-50 | 49 (10.7) | 32 (9.1) | 60 (10.7) | 17 (3.7) | 133 (10.9) |
| 51-60 | 92 (20.0) | 62 (17.7) | 90 (16.1) | 65 (14.1) | 242 (19.8) |
| 61-70 | 110 (23.9) | 71 (20.2) | 123 (21.9) | 127 (27.6) | 320 (26.1) |
| 71-80 | 108 (23.5) | 73 (20.8) | 120 (21.4) | 121 (26.3) | 202 (16.5) |
| 81-90 | 64 (13.9) | 51 (14.5) | 101 (18.0) | 92 (20) | 102 (8.3) |
| At least 90 | 10 (2.2) | 8 (2.3) | 27 (4.8) | 27 (5.9) | 31 (2.5) |
| **Previous Medical History, n (%)** |  |  |  |  |  |
| Hypertension (%) | 228 (49.7) | 120 (34.2) | 210 (37.5) | 128 (27.8) | 462 (37.7) |
| Atrial Fibrillation (%) | 45 (9.8) | 18 (5.1) | 40 (7.1) | 27 (5.9) | 86 (7.0) |
| Coronary Artery Disease (%) | 95 (20.7) | 46 (13.1) | 85 (15.2) | 81 (17.6) | 171 (13.9) |
| Heart failure (%) | 51 (11.1) | 27 (7.7) | 61 (10.9) | 42 (9.1) | 110 (8.9) |
| Stroke (%) | 43 (9.4) | 22 (6.3) | 39 (6.9) | 14 (3.0) | 98 (8) |
| Chronic Kidney Disease (%) | 54 (11.8) | 20 (5.7) | 58 (10.4) | 26 (5.6) | 136 (11.1) |
| Diabetes (%) | 154 (33.6) | 68 (19.4) | 143 (25.5) | 101 (21.9) | 313 (25.6) |
| Asthma (%) | 42 (9.2) | 32 (9.1) | 45 (8.0) | 13 (2.8) | 115 (9.4) |
| Chronic Obstructive Pulmonary Disease (%) | 35 (7.6) | 16 (4.6) | 31 (5.5) | 21 (4.6) | 65 (5.1) |
| Cancer (%) | 20 (4.4) | 30 (8.5) | 33 (5.9) | 11 (2.4) | 112 (9.1) |
| **Vital Signs at Hospital Admission, Median (IQR)** |  |  |  |  |  |
| Heart Rate (bpm) | 89<br>(78 – 101) | 89<br>(78-100.2) | 89<br>(78-100) | 85<br>(76-96) | 89<br>(79-101.3) |
| Oxygen Saturation (%) | 95<br>(93-97) | 96<br>(94-98) | 96<br>(94-98) | 96<br>(94.5-98) | 96<br>(94-98) |
| Respiration Rate (breaths / minute) | 18<br>(18-20) | 18<br>(18-19.3) | 20<br>(18-22) | 18<br>(17- 20) | 20<br>(18-22) |
| Temperature (F) | 98.8<br>(98.2-99.7) | 98<br>(97.3-98.7) | 98.7<br>(98.1-99.9) | 98.7<br>(98-99.6) | 99<br>(98.2-100) |
| Systolic Blood Pressure (mmHg) | 70<br>(63-78) | 70<br>(62.75-80) | 69<br>(61-78) | 74<br>(65.5-84) | 74<br>(66.8-82) |
| Diastolic Blood Pressure (mmHg) | 123<br>(110-138) | 130<br>(112-145) | 124<br>(112-138) | 126<br>(111-137) | 128.5<br>(114-144) |
| Weight (kg) | 76.3<br>(65.2-90.7) | 79.4<br>(68.1-92.9) | 80.9<br>(69.0-95.3) | 78.8<br>(67.6-90.7) | 79.6<br>(69.1-92.2) |
| **Admission Laboratory Parameters, Median (IQR)** |  |  |  |  |  |
| **Metabolic Markers** |  |  |  |  |  |
| Sodium (mEq/L) | 138<br>(136-141) | 139<br>(137-142) | 137<br>(135-140) | 137<br>(134-139) | 138<br>(135-141) |
| Potassium (mEq/L) | 4.3<br>(3.9-4.7) | 4.3<br>(3.8-4.7) | 4<br>(3.6-4.5) | 4.2<br>(3.9-4.7) | 4.2<br>(3.8-4.7) |
| Creatinine (mg/dL) | 1<br>(0.8-2.1) | 1.2<br>(0.9-2.1) | 0.9<br>(0.7-1.4) | 0.9<br>(0.7-1.3) | 1.0<br>(0.8-1.7) |
| Lactate (mg/dL) | - | 1.94<br>(1.1-2.9) | 1.7<br>(1.3-2.2) | 1.5<br>(1.1-2) | 1.3<br>(1-1.8) |
| **Hematological Markers** |  |  |  |  |  |
| White Blood Cells (103/μL) | 7.9<br>(5.9-10.5) | 7.5<br>(5.4-10.5) | 6.8<br>(5.2-9.8) | 7.7<br>(5.6-10.9) | 7.5<br>(5.7-10.5) |
| Lymphocyte Percentage | 12<br>(7.1-18.3) | 14.7<br>(8.9-22.7) | 13.1<br>(7.8-20.1) | 16.1<br>(9.6-23.1) | 13.05<br>(9-20.5) |
| Hemoglobin (mEq/L) | 12.6<br>(11.1-13.8) | 12.5<br>(11.2-13.8) | 12.5<br>(11.1-13.7) | 13<br>(11.6-14.2) | 13<br>(11.6-14) |
| Red Blood Cell Distribution Width (%) | 12.8<br>(12.1-14.2) | 13.1<br>(12.5-14.1) | 14.4<br>(13.6-15.6) | 12.5<br>(12-13.5) | 12.6<br>(12.1-13.7) |
| Platelets (#) | 188<br>(146-259) | 205.5<br>(153-266.2) | 212<br>(162-279.3) | 197<br>(152-264) | 187<br>(144-252) |
| **Liver Function** |  |  |  |  |  |
| Alanine Aminotransferase (units/L) | 34<br>(21-55) | 28<br>(19-49) | 31<br>(19-54) | 30<br>(19-54) | 32<br>(19-54.8) |
| Aspartate Aminotransferase (units/L) | 50<br>(34-76.8) | 42<br>(28-74) | 41.5<br>(29-67) | 48.5<br>(32-77) | 45<br>(31-71) |
| Albumin (g/dL) | 2.7<br>(2.3-3.2) | 3.4<br>(3.1-3.7) | 3<br>(2.6-3.3) | 3<br>(2.6-3.3) | 2.9<br>(2.5-3.1) |
| Total Bilirubin (mg/dL) | 0.6<br>(0.4-0.7) | 0.6<br>(0.4-0.7) | 0.6<br>(0.4-0.8) | 0.5<br>(0.4-0.8) | 0.6<br>(0.4-0.8) |
| **Coagulation Markers** |  |  |  |  |  |
| Prothrombin Time (s) | 14<br>(13.4-17.2) | 14.6<br>(13.5-16.1) | 14.3<br>(13.5-15.7) | 13.9<br>(13.4-14.9) | 14.2<br>(13.6-15.3) |
| Partial Thromboplastin Time (s) | 35.7<br>(32.2-61.2) | 35.2<br>(32.3-38.6) | 32.5<br>(29.2-37.2) | 32.1<br>(29.8-37.5) | 32.9<br>(29.4-38.7) |
| **Gases** |  |  |  |  |  |
| PCO2 (mmHg) | - | 40<br>(34.3-46) | 41<br>(37-47) | 41.5<br>(36.3-48) | 41<br>(35-47) |
| pH | - | 7.4<br>(7.3-7.4) | 7.4<br>(7.4-7.4) | 7.4<br>(7.3-7.4) | 7.4<br>(7.3-7.4) |
| **Inflammatory Markers** |  |  |  |  |  |
| C Reactive Protein (mg/L) | 207.2<br>(159.4-233.4) | 147.7<br>(90.7-243.3) | 152.3<br>(83.1-205.6) | 104.9<br>(50.6-190.4) | 125.4<br>(62.7-201.6) |
| Ferritin (ng/mL) | 968<br>(473.6-2378.3) | 999.5<br>(522.2-2228.5) | 720<br>(343.5-1813.8) | 713.5<br>(305.8-1466) | 763<br>(360- 1784) |
| D-Dimer (ng/mL) | 1.5<br>(1.0-2.9) | 2.3<br>(1.1-4.1) | 1.3<br>(0.7-2.5) | 1.4<br>(0.7-2.6) | 1.5<br>(0.9-2.9) |
| Creatinine Phosphokinase (units/L) | 237.5<br>(187.8-440.5) | - | 156<br>(80-505) | - | - |
| Procalcitonin (ng/mL) | 0.3<br>(0.1-0.8) | 0.4<br>(0.2-1.4) | 0.2<br>(0.1-0.6) | 0.2<br>(0.1-0.5) | 0.2<br>(0.1-0.7) |
| Lactate Dehydrogenase (units/L) | 468<br>(359.5-628) | 489<br>(390-637.5) | 417<br>(315-559) | 448<br>(345-651.5) | 427<br>(324-559) |
| **Cardiac Markers** |  |  |  |  |  |
| Troponin I (ng/mL) | 0.2<br>(0.1-0.9) | - | 0.1<br>(0.0-0.2) | - | - |
| **Outcomes** |  |  |  |  |  |
| Critical | 0<br>(0-1) | 0<br>(0-1) | 0<br>(0-1) | 0<br>(0-1) | 0<br>(0-1) |
| Mortality | 0<br>(0-0) | 0<br>(0-0) | 0<br>(0-0) | 0<br>(0-0) | 0<br>(0-0) |

**Supplementary Table 2.**
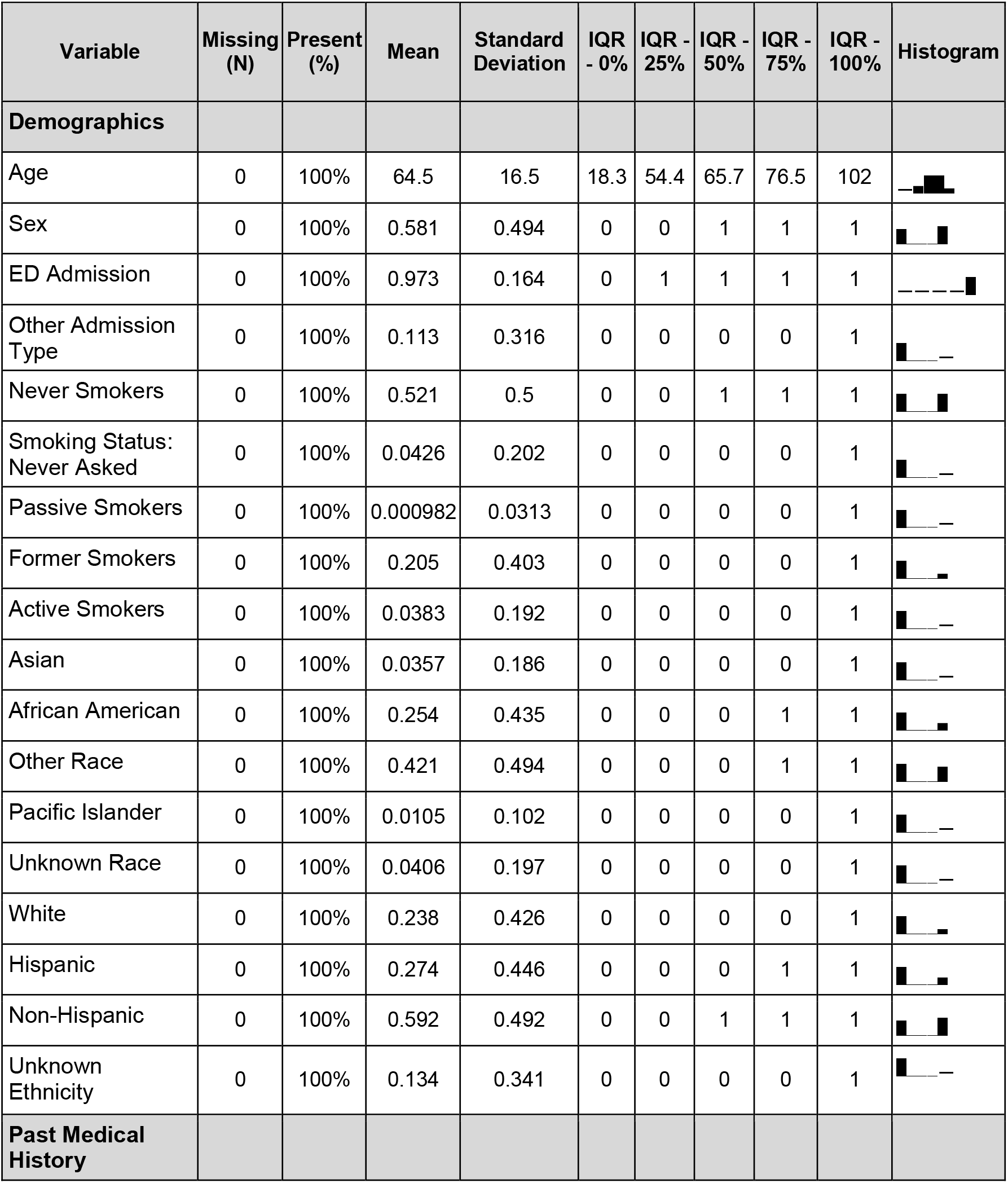
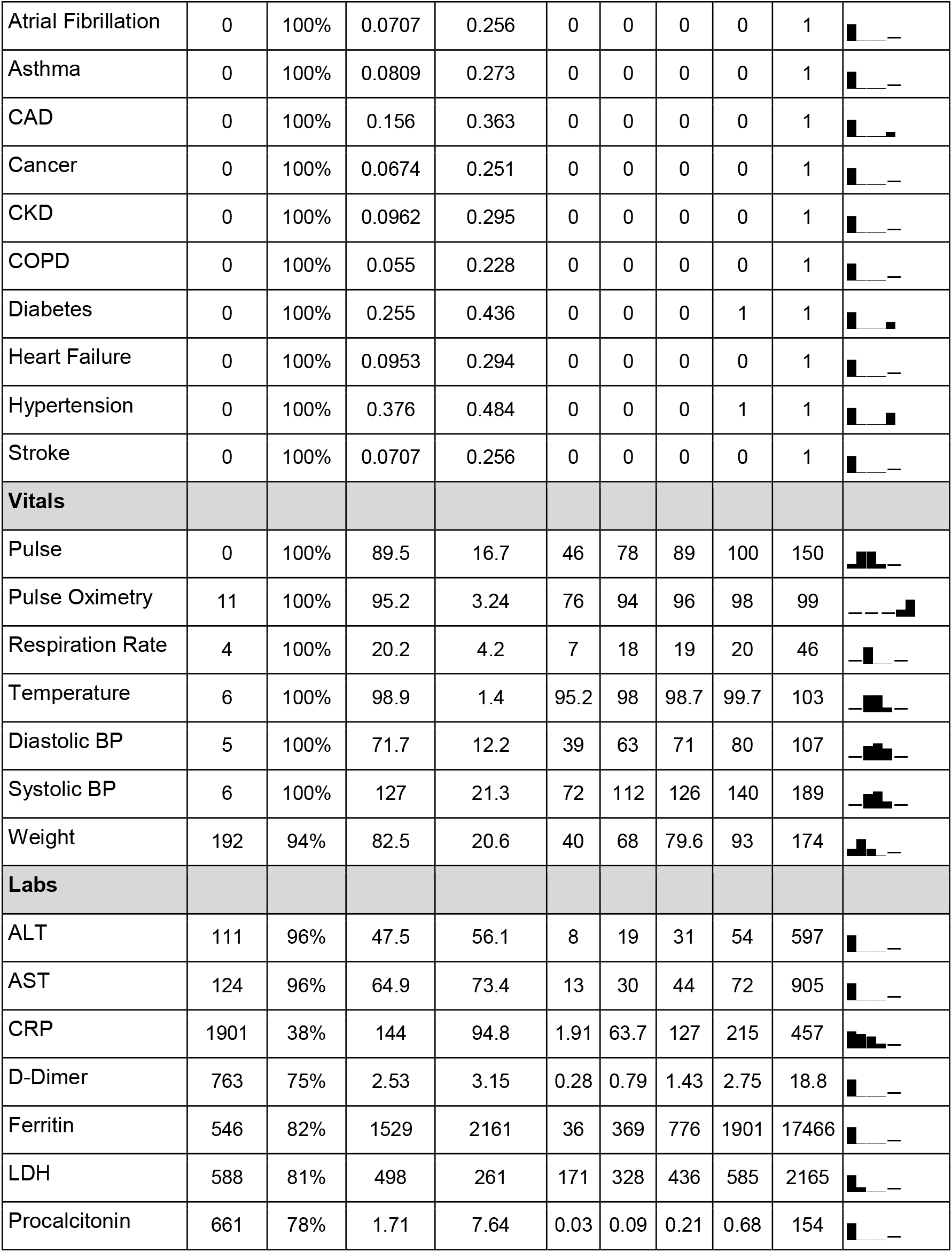
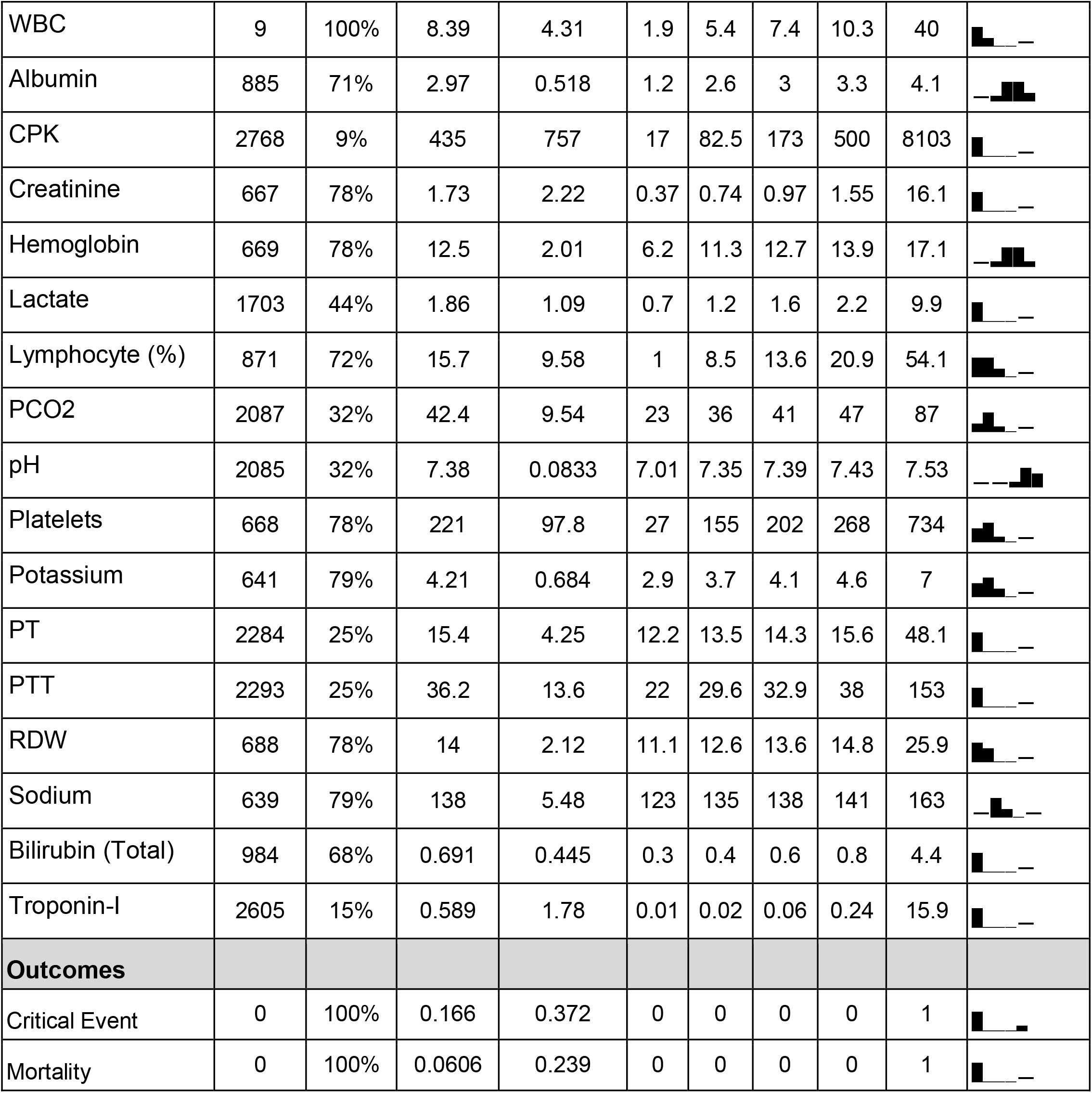
Baseline Patient Feature Analysis. Missingness, means, standard deviations, interquartile ranges, and histograms for all features in the dataset.

**Supplementary Table 3.**
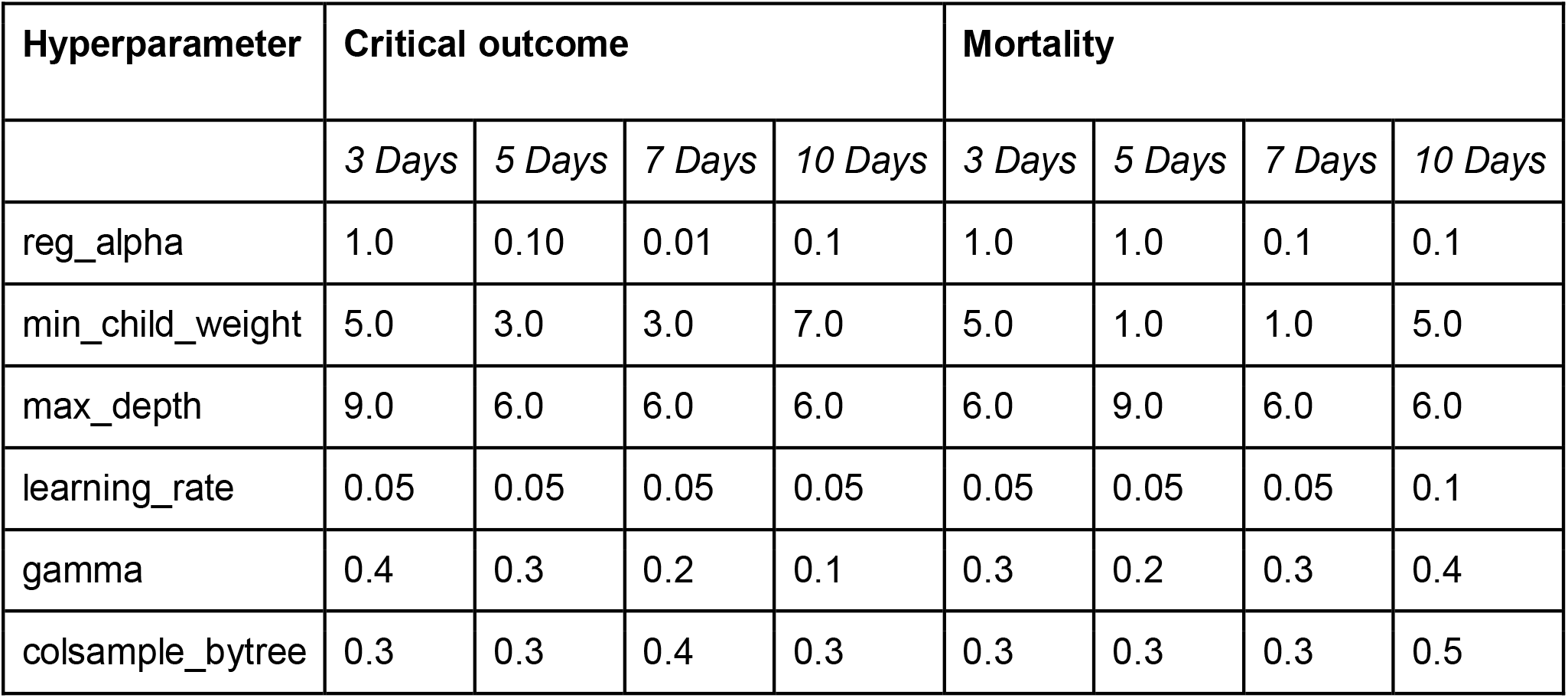
Final XGBoost Model Hyperparameters. Final hyperparameters for the XGBoost classifier at day 7 for critical event and mortality prediction after tuning using a grid-search to optimize for AUC-ROC.

**Supplementary Table 4.** Model Performance Across Each Hospital. Performance of the XGBoost classifier by hospital site, as measured by accuracy (ACC), area under the receiver operating curve (AUCROC), area under the precision recall curve (AUCPRC), F1-score (F1), sensitivity (SENS), and specificity (SPEC). “OH” refers to all hospitals in the external validation set (i.e. MSW, MSM, MSB, MSQ). Outcomes are structured by “<outcome>_<day>“, where outcome is either a critical event (CRITICAL) or mortality (MORTALITY) and the time frame it was predicted over. “OUTCOME PROP” refers to the portion of the dataset with the respective outcome.

| FACILITY | OUTCOME | PATIENTS | PROP<br>OUTCOME | ACC | AUCROC | AUCPRC | F1 | SENS | SPEC |
| --- | --- | --- | --- | --- | --- | --- | --- | --- | --- |
| MSH | CRITICAL_3 | 1225 | 0.17 | 0.90 | 0.89 | 0.71 | 0.59 | 0.84 | 0.90 |
|  | CRITICAL_5 | 1156 | 0.22 | 0.84 | 0.84 | 0.67 | 0.53 | 0.75 | 0.85 |
|  | CRITICAL_7 | 1052 | 0.25 | 0.81 | 0.83 | 0.66 | 0.53 | 0.71 | 0.83 |
|  | CRITICAL_10 | 931 | 0.28 | 0.80 | 0.83 | 0.67 | 0.57 | 0.70 | 0.82 |
|  | MORTALITY_3 | 1225 | 0.03 | 0.97 | 0.87 | 0.26 | 0.03 | 1.00 | 0.97 |
|  | MORTALITY_5 | 1156 | 0.05 | 0.95 | 0.84 | 0.37 | 0.13 | 0.56 | 0.95 |
|  | MORTALITY_7 | 1052 | 0.07 | 0.93 | 0.87 | 0.49 | 0.21 | 0.71 | 0.94 |
|  | MORTALITY_10 | 931 | 0.12 | 0.90 | 0.84 | 0.50 | 0.36 | 0.65 | 0.91 |
| MSQ | CRITICAL_3 | 459 | 0.20 | 0.81 | 0.79 | 0.52 | 0.21 | 0.60 | 0.82 |
|  | CRITICAL_5 | 440 | 0.29 | 0.73 | 0.79 | 0.59 | 0.21 | 0.67 | 0.73 |
|  | CRITICAL_7 | 414 | 0.36 | 0.69 | 0.77 | 0.65 | 0.31 | 0.71 | 0.68 |
|  | CRITICAL_10 | 390 | 0.41 | 0.69 | 0.80 | 0.73 | 0.46 | 0.80 | 0.67 |
|  | MORTALITY_3 | 459 | 0.14 | 0.86 | 0.77 | 0.42 | 0.00 | - | 0.86 |
|  | MORTALITY_5 | 440 | 0.23 | 0.80 | 0.76 | 0.56 | 0.21 | 0.92 | 0.79 |
|  | MORTALITY_7 | 414 | 0.30 | 0.74 | 0.77 | 0.61 | 0.25 | 0.86 | 0.73 |
|  | MORTALITY_10 | 390 | 0.37 | 0.68 | 0.78 | 0.67 | 0.29 | 0.74 | 0.67 |
| MSB | CRITICAL_3 | 460 | 0.17 | 0.85 | 0.77 | 0.45 | 0.20 | 0.75 | 0.85 |
|  | CRITICAL_5 | 426 | 0.28 | 0.75 | 0.80 | 0.63 | 0.19 | 0.93 | 0.74 |
|  | CRITICAL_7 | 397 | 0.35 | 0.68 | 0.76 | 0.61 | 0.17 | 0.81 | 0.67 |
|  | CRITICAL_10 | 370 | 0.41 | 0.65 | 0.80 | 0.71 | 0.32 | 0.79 | 0.63 |
|  | MORTALITY_3 | 460 | 0.10 | 0.90 | 0.79 | 0.39 | 0.04 | 1.00 | 0.90 |
|  | MORTALITY_5 | 426 | 0.21 | 0.80 | 0.83 | 0.58 | 0.11 | 1.00 | 0.80 |
|  | MORTALITY_7 | 397 | 0.28 | 0.75 | 0.82 | 0.66 | 0.22 | 1.00 | 0.74 |
|  | MORTALITY_10 | 370 | 0.36 | 0.67 | 0.79 | 0.67 | 0.19 | 0.88 | 0.66 |
| MSW | CRITICAL_3 | 351 | 0.12 | 0.89 | 0.81 | 0.46 | 0.31 | 0.53 | 0.90 |
|  | CRITICAL_5 | 331 | 0.15 | 0.86 | 0.78 | 0.45 | 0.43 | 0.57 | 0.89 |
|  | CRITICAL_7 | 307 | 0.16 | 0.87 | 0.81 | 0.52 | 0.44 | 0.64 | 0.89 |
|  | CRITICAL_10 | 280 | 0.20 | 0.83 | 0.80 | 0.58 | 0.49 | 0.59 | 0.87 |
|  | MORTALITY_3 | 351 | 0.02 | 0.98 | 0.89 | 0.09 | 0.00 | - | 0.98 |
|  | MORTALITY_5 | 331 | 0.03 | 0.97 | 0.86 | 0.23 | 0.00 | - | 0.97 |
|  | MORTALITY_7 | 307 | 0.05 | 0.95 | 0.88 | 0.38 | 0.00 | 0.00 | 0.95 |
|  | MORTALITY_10 | 280 | 0.07 | 0.95 | 0.80 | 0.41 | 0.40 | 1.00 | 0.95 |
| **MSM** | CRITICAL_3 | 560 | 0.16 | 0.82 | 0.73 | 0.34 | 0.20 | 0.36 | 0.85 |
|  | CRITICAL_5 | 525 | 0.22 | 0.78 | 0.73 | 0.41 | 0.30 | 0.50 | 0.81 |
|  | CRITICAL_7 | 481 | 0.26 | 0.76 | 0.75 | 0.50 | 0.34 | 0.63 | 0.78 |
|  | CRITICAL_10 | 420 | 0.32 | 0.75 | 0.78 | 0.64 | 0.50 | 0.68 | 0.76 |
|  | MORTALITY_3 | 560 | 0.06 | 0.94 | 0.89 | 0.43 | 0.00 | - | 0.94 |
|  | MORTALITY_5 | 525 | 0.10 | 0.90 | 0.87 | 0.48 | 0.04 | 1.00 | 0.90 |
|  | MORTALITY_7 | 481 | 0.14 | 0.87 | 0.84 | 0.49 | 0.14 | 1.00 | 0.87 |
|  | MORTALITY_10 | 420 | 0.20 | 0.84 | 0.84 | 0.61 | 0.34 | 0.82 | 0.84 |
| **OH** | CRITICAL_3 | 1830 | 0.17 | 0.84 | 0.77 | 0.43 | 0.23 | 0.57 | 0.85 |
|  | CRITICAL_5 | 1722 | 0.24 | 0.78 | 0.76 | 0.51 | 0.26 | 0.65 | 0.79 |
|  | CRITICAL_7 | 1599 | 0.29 | 0.73 | 0.74 | 0.52 | 0.30 | 0.60 | 0.75 |
|  | CRITICAL_10 | 1460 | 0.34 | 0.73 | 0.79 | 0.66 | 0.44 | 0.77 | 0.73 |
|  | MORTALITY_3 | 1830 | 0.08 | 0.92 | 0.83 | 0.38 | 0.03 | 1.00 | 0.92 |
|  | MORTALITY_5 | 1722 | 0.15 | 0.86 | 0.82 | 0.52 | 0.08 | 1.00 | 0.86 |
|  | MORTALITY_7 | 1599 | 0.20 | 0.82 | 0.84 | 0.58 | 0.18 | 0.86 | 0.82 |
|  | MORTALITY_10 | 1460 | 0.26 | 0.79 | 0.83 | 0.64 | 0.40 | 0.74 | 0.79 |

**Supplementary Table 5.** Model Performance At Extended Time Frames. Performance of the XGBoost classifier on validation set from MSH and external validation set (MSB, MSW, MSQ, MSM), as measured by accuracy (ACC), area under the receiver operating curve (AUCROC), area under the precision recall curve (AUCPRC), F1-score (F1), sensitivity (SENS), and specificity (SPEC). “OH” refers to all hospitals in the external validation set (i.e. MSW, MSM, MSB, MSQ). Outcomes are structured by “<outcome>_<day>“, where outcome is either a critical event (CRITICAL) or mortality (MORTALITY) and the time frame it was predicted over. “OUTCOME PROP” refers to the portion of the dataset with the respective outcome.

| FACILITY | OUTCOME | PATIENTS | PROP<br>OUTCOME | ACC | AUCROC | AUCPRC | F1 | SENS | SPEC |
| --- | --- | --- | --- | --- | --- | --- | --- | --- | --- |
| MSH | CRITICAL_15 | 792 | 0.27 | 0.81 | 0.84 | 0.68 | 0.60 | 0.71 | 0.84 |
|  | CRITICAL_20 | 719 | 0.25 | 0.82 | 0.85 | 0.68 | 0.58 | 0.70 | 0.85 |
|  | MORTALITY_15 | 792 | 0.16 | 0.87 | 0.86 | 0.58 | 0.58 | 0.64 | 0.89 |
|  | MORTALITY_20 | 719 | 0.18 | 0.86 | 0.87 | 0.64 | 0.54 | 0.67 | 0.87 |
| OH | CRITICAL_15 | 1320 | 0.37 | 0.72 | 0.75 | 0.66 | 0.53 | 0.68 | 0.73 |
|  | CRITICAL_20 | 1254 | 0.36 | 0.75 | 0.81 | 0.71 | 0.57 | 0.75 | 0.75 |
|  | MORTALITY_15 | 1320 | 0.32 | 0.74 | 0.81 | 0.65 | 0.40 | 0.75 | 0.74 |
|  | MORTALITY_20 | 1254 | 0.34 | 0.75 | 0.84 | 0.72 | 0.51 | 0.77 | 0.75 |

**Supplementary Table 6.** Logistic Regression and Generalized Additive Model Performance. Performance of the four models using only Age and all the features, as measured by accuracy (ACC), area under the receiver operating curve (AUCROC), area under the precision recall curve (AUCPRC), F1-score (F1), sensitivity (SENS), and specificity (SPEC). Models are structured by “<model>_<feature space>“, where model is either a logistic regression (LOG) or generalized additive model (GAM) and the features used in the model.

| MODEL | OUTCOME | PATIENTS | PROP<br>OUTCOME | ACC | AUCROC | AUCPRC | F1 | SENS | SPEC |
| --- | --- | --- | --- | --- | --- | --- | --- | --- | --- |
| LOG_AGE | CRITICAL_3 | 3055 | 0.17 | 0.83 | 0.57 | 0.22 | 0 | 1 | 0 |
|  | MORTALITY_3 | 3055 | 0.06 | 0.92 | 0.70 | 0.18 | 0 | 1 | 0 |
| GAM_AGE | CRITICAL_3 | 3055 | 0.17 | 0.83 | 0.52 | 0.19 | 0.01 | 1 | 0.003 |
|  | MORTALITY_3 | 3055 | 0.06 | 0.98 | 0.62 | 0.14 | 0 | 1 | 0 |
| LOG_ALL | CRITICAL_3 | 1278 | 0.18 | 0.81 | 0.74 | 0.33 | 0.31 | 0.90 | 0.29 |
|  | MORTALITY_3 | 1278 | 0.02 | 0.96 | 0.67 | 0.06 | 0.07 | 0.98 | 0.07 |
| GAM_ALL | CRITICAL_3 | 1278 | 0.18 | 0.82 | 0.74 | 0.37 | 0.35 | 0.92 | 0.31 |
|  | MORTALITY_3 | 1278 | 0.02 | 0.98 | 0.62 | 0.14 | 0 | 1 | 0 |

